# Morbidity, Mortality and Immunization in Severe COVID-19 Among the Elderly in Brazil: A Five-Year Perspective

**DOI:** 10.1101/2025.05.01.25326813

**Authors:** Silvio Alencar Cândido-Sobrinho, Francisco de Sousa Júnior, José Quirino da Silva-Filho, Vânia Angélica Feitosa Viana, Aldo Ângelo Moreira Lima

## Abstract

The COVID-19 pandemic underscored longstanding vulnerabilities within Brazil’s healthcare system, particularly affecting the elderly population in underserved regions. Although infection can affect individuals across all age groups, the elderly population is particularly susceptible to severe outcomes due to aging-related factors. Despite the availability of effective vaccines, full immunization coverage was not achieved, allowing the progression of cases to severe acute respiratory syndrome (SARS). This study aimed to analyse the evolution of severe COVID-19 cases in parallel with mild and moderate cases among the Brazilian elderly population between January 2020 and December 2024, spanning five years, while considering regional and social differences, circulating variants, vaccination, and factors associated with COVID-19-related SARS in Brazil. A total of 15,609 cases of influenza-like illness (mild and moderate) and 580,818 PCR-confirmed cases were recorded, of which 304,341 resulted in recovery and 276,477 in death, with a case fatality rate of 47.60%. Brazil experienced three waves of COVID-19-related SARS, with the second wave being the most lethal across all regions. The Gamma and Omicron variants were the most persistent and impactful. The transition between variants influenced the regional dynamics of the pandemic, although little variation was observed in the proportion of circulating variants across regions. The study highlights the importance of continuous monitoring, genomic surveillance, and vaccination coverage to anticipate and mitigate future pandemic waves.

**WHAT WAS ALREADY KNOWN?:**

- **Elderly individuals are more vulnerable to COVID-19:** Previous studies had already shown that aging and the presence of comorbidities increased the severity of COVID-19 in this group.
- **Regional inequalities influence outcomes:** Regions with lower Human Development Index (HDI), limited access to healthcare, and greater social vulnerability presented worse mortality indicators.
- **Vaccination reduces mortality:** It was already known that complete vaccination, especially with booster doses, significantly reduced the severity of COVID-19.

**WHAT IS NEW?:**

- **More detailed epidemiological profile by region and year:** The study traces the pattern of lethality over a five-year period (2020-2024), revealing significant fluctuations across regions, age groups, educational levels, ethnicities, and geographic zones.
- **High impact of multimorbidity:** The article shows that elderly individuals with three or more comorbidities had the lowest survival rates and the highest risk of death, regardless of their vaccination status.
- **Clear protective effect of booster doses:** The Kaplan-Meier survival analysis demonstrated that only individuals who received a first and/or second booster showed a significant increase in median survival time.

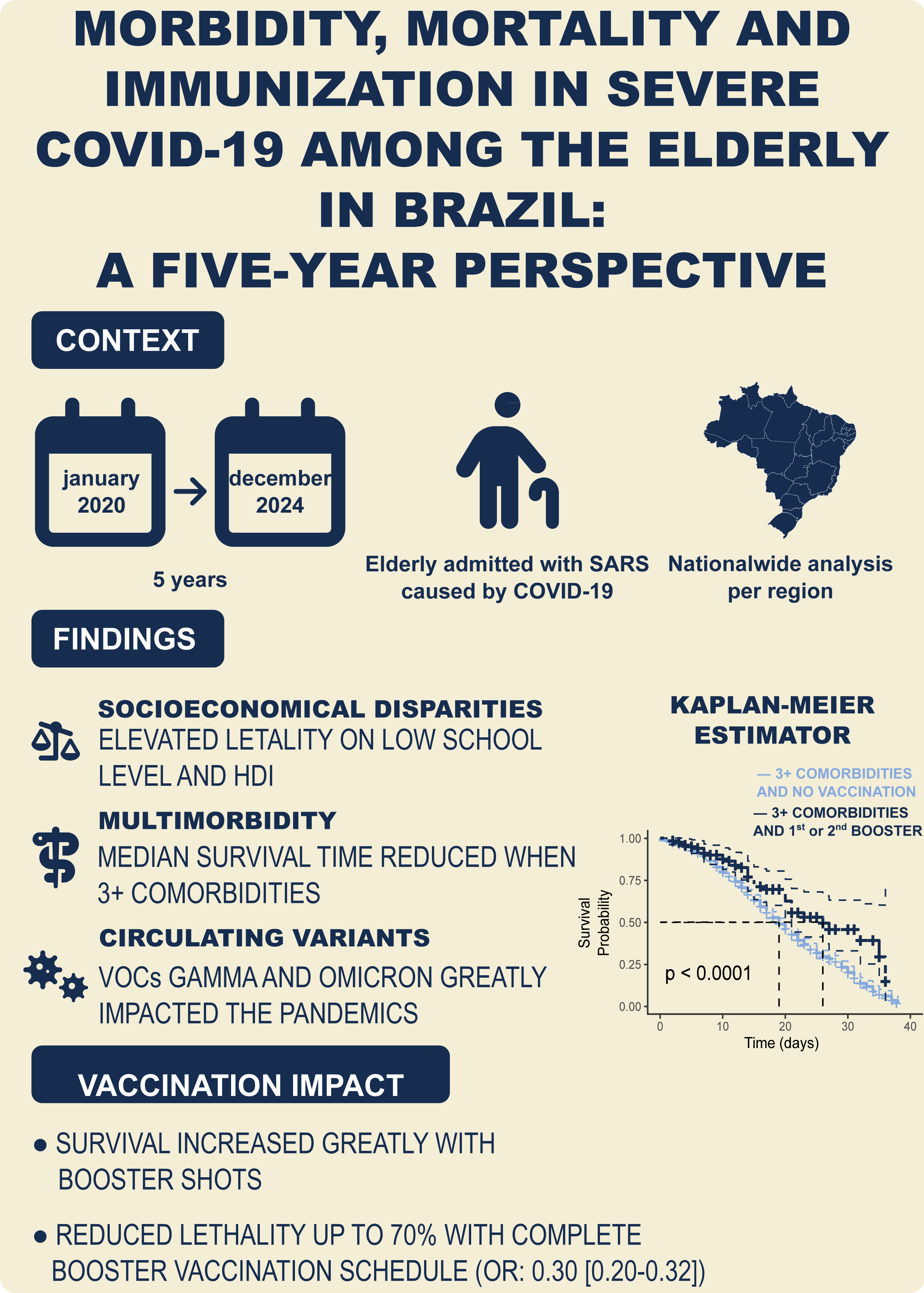

## INTRODUCTION

In March 2020, the World Health Organization (WHO) declared a state of pandemic due to the rapid global spread of the SARS-CoV-2 virus, identified a few months earlier in China^1^. This event posed numerous challenges to healthcare systems worldwide, having a particularly severe impact on developing countries and those with fragile healthcare system^2^.

SARS-CoV-2 can cause severe infections requiring hospitalization and intensive care due to its high transmissibility rate and the severity of respiratory complications, leading to the overload of healthcare systems globally and in Brazil^3–5^.

Although the infection can occur across all age groups, with clinical manifestations ranging from asymptomatic cases to death, elderly individuals have been shown to be more susceptible to complications and fatal outcomes. This increased vulnerability is associated with aging-related factors, such as decreased immune system efficiency and a higher prevalence of multimorbidity^6^. Moreover, sociodemographic determinants such as region, geographic setting, age group, and sex at birth, alongside socioeconomic factors like the Human Development Index (HDI), race/ethnicity, and educational attainment, may influence survival outcomes across all age groups. Identifying which factors most adversely affect the elderly population is critical^7^.

Following the identification of the etiological agent and the sequencing of its genome in 2020, institutions began studies to develop vaccines. By early 2021, some vaccines had been approved for emergency use and subsequently for full regulatory authorization in Brazil. Due to the increased mortality risk associated with older age^8^, the elderly were prioritized in the national immunization campaign. However, challenges such as vaccine hesitancy and the emergence of new viral variants compromised vaccination uptake and raised concerns regarding the effectiveness of the immunization efforts at that time^9^. Given the heightened vulnerability of the elderly to severe COVID-19 outcomes, a detailed analysis of the vaccination’s impact on survival in this population becomes essential.

Thus, this study aimed to describe the dynamics of COVID-19 among the elderly population in Brazil, considering regional and social disparities, multimorbidity, the impact of circulating variants, vaccination, and key factors associated with progression to severe acute respiratory syndrome (SARS).

## METHODS

### Research Ethics

This study used aggregated data without the possibility of individual identification, made available in public repositories, and was therefore exempt from evaluation by a Research Ethics Committee according to the Brazilian National Health Council Resolution No. 510/2016^10^.

### Study Design

This is an observational study of the retrospective cohort^11^, based on secondary data extracted from national health surveillance systems, encompassing mild, moderate, and severe cases of COVID-19 in the Brazilian elderly population between 2020 and 2024.

### Data Source

Data covering the period from January 1, 2020, to December 31, 2024, were retrieved as follows: mild and moderate cases were obtained from the ‘Influenza-like Illness Notifications Database’ (referred as ‘ILI’); severe cases were obtained from the ‘Severe Acute Respiratory Syndrome Database - including COVID-19 data’ (referred as ‘SARS’); and Vaccination data were obtained from the ‘National COVID-19 Vaccination Campaign Database’, all retrieved from openDataSUS repositories^12^. Information on circulating variants of SARS-CoV-2 in Brazil during the study period was retrieved from the Fiocruz Genomic Network repositories^13^. Human Development Index (HDI) 2010 data by municipality were obtained from the United Nations Development Programme^14^. Population data were retrieved from the IBGE Automatic Recovery System using *sidrar*^15^.

### Inclusion and Exclusion Criteria

From the ILI database (mild and moderate cases), Individuals aged between 60 and 112 years, regardless of sex or region, with laboratory-confirmed COVID-19. Included cases were those that evolved to cure, death, or home treatment. Individuals outside the 60-112 age range, or those whose records lacked information on state of residence, notification date, or etiologic agent (unspecified SARS or suspected cases), were excluded.

Regarding the SARS database (severe cases), individuals aged between 60 and 112 years with SARS-CoV-2 confirmation by PCR were included if their final case classification was either ‘Cure’ or ‘Death,’ with an length of stay between 0 and 38 days (IQR = 15.85 [10-20] days), and with vaccination records in sequential order (‘None’, 1^st^ Dose’, ‘2^nd^ Dose’, ‘1^st^ Booster’, and ‘2^nd^ Booster’), including manufacturer information and a minimum interval of seven days after vaccination. Individuals without PCR confirmation, aged below 60 or above 112 years, those whose deaths were unrelated to COVID-19, or cases lacking outcome information (ignored cases), as well as those with outlier length of stay or vaccination data, were excluded.

Regarding the National COVID-19 Vaccination Campaign data, individuals of any sex or region, aging from 60 to 112 years who received the 1^st^ Dose, 2^nd^ Dose, 1^st^ Booster, or 2^nd^ Booster were included. Individuals for whom age or state of residence could not be identified, as well as records of doses related to exceptional cases or new vaccination schedules were excluded.

### Data Processing and Statistical Analysis

The data were processed using AWK, bash, and R^16^ programming languages, with the *tidyverse* libraries used for data processing and string detection^17^. Graphs were generated with *ggplot2*^18^, *ggpubr*^19^, *survminer*^20^ and *forestmodel*^21^ libraries. Contingency tables were created using *tableone*^22^.

Variables were expressed as mean ± standard deviation or median [IQR1-IQR3]. Adjusted incidence and mortality rates were calculated based on data from the 2022 Census. Differences in proportions were assessed using the chi-square test (χ²), and comparisons of case fatality rates (CFR) between groups were performed using the Z-Test for the comparison of multiple two-sided proportions with p-value adjustment by the Bonferroni method using *rstatix*^23^ and *rcompanion*^24^. Analysis of Variance (ANOVA) was performed using the *stats* library, and Tukey’s test was conducted using *agricolae*^25^ library.

Odds Ratios (OR) were expressed as OR [lower 95% CI - upper 95% CI] and were estimated using Generalized Linear Models (GLM). For this analysis, only complete cases (n = 178,058) were considered. Univariate models were adjusted using 73 variables, including demographic characteristics (Region, Age Group, HDI 2010, Sex, Ethnicity/Race, Education Level, and Geographic Zone); signs and symptoms (General: fever, chills, malaise, sweating, asthenia, prostration, fatigue, loss of appetite, hyporexia, insomnia; Respiratory: cough, runny nose, bronchitis, sneezing, sore throat, wheezing, tachypnea, respiratory failure, chest pain, cyanosis, hemoptysis, oxygen saturation SpO2 < 95%; Neurological: headache, confusion, seizure; Musculoskeletal: myalgia, arthralgia, back pain; Gastrointestinal: nausea, vomiting, abdominal pain, diarrhea; and Other: tachycardia, edema, conjunctivitis, loss of smell, and loss of taste); and risk factors (asthma, heart disease, lung disease, chronic kidney disease, immunosuppression, chronic liver disease, neurological disorders, obesity, postpartum status, Down syndrome, diabetes, Alzheimer’s disease, pancreatitis, alcoholism, anemia, COPD, stroke, hypertension, emphysema, hypothyroidism, arthritis, ascites, lymphoma, lupus, and cancer). Variables with p-values < 0.05 and OR > 1.0 in univariate models were selected for multivariate modelling, resulting in a total of 29 variables. The final model was fitted using forward-backward stepwise regression based on the MASS library^26^.

For modelling purposes, individuals were categorized according to their vaccination status into three groups: those who received no vaccination, those who received the first and/or second dose (‘1^st^/2^nd^ Dose’), and those who received the first and/or second booster (‘1^st^/2^nd^ Booster’). Additionally, the number of comorbidities/risk factors per individual was calculated, including diabetes, neuropathy, chronic kidney disease, chronic lung disease, immunosuppression, and obesity. These risk factors were selected based on a GLM model adjusted by stepwise selection. Individuals were then classified into three groups according to the total number of reported comorbidities/risk factors: ‘0’ (no comorbidities/risk factors), ‘1-2’ (one or two comorbidities/risk factors), and ‘3+’ (three or more comorbidities/risk factors).

Finally, the effect of vaccination status combined with the number of comorbidities/risk factors on median survival time was evaluated through multivariate adjustment of Kaplan-Meier survival curves in the population of severe cases. Comparisons between survival curves were validated using the log-rank test, employing the *survival*^27^ and *survminer*^20^ libraries.

## RESULTS

During the study period (January 1, 2020, to December 31, 2024), a total of 24,970 cases of Influenza-like Illness (ILI) were reported among older adults (aged 60 to 112 years), of which 15,609 had laboratory confirmation. In contrast, during the same period, 4,029,767 cases of Severe Acute Respiratory Syndrome (SARS) were recorded, with SARS-CoV-2 identified as the etiological agent in 2,127,427 cases, and 1,171,801 confirmed by PCR testing (Figure S1). Among the PCR-confirmed cases, 580,818 were recorded for the elderly population aged 60 to 112 years, representing 49.6% of all PCR-confirmed COVID-19 SARS cases in Brazil (Figure S1). Immunization among the elderly during this period included 30,514,826 1^st^ Doses, 29,823,907 2^nd^ Doses, followed by 41,675,236 1^st^ Booster doses, and 16,732,112 2^nd^ Boosters.

The onset of the pandemic was marked by a surge in cases and deaths, with well-defined incidence and mortality waves observed across Brazil (Figure 1). The incidence and mortality rates for COVID-19-related SARS revealed three major waves, each characterized by sharp peaks and distinct regional fluctuations (Figure 2A-J).

**Figure 1.**
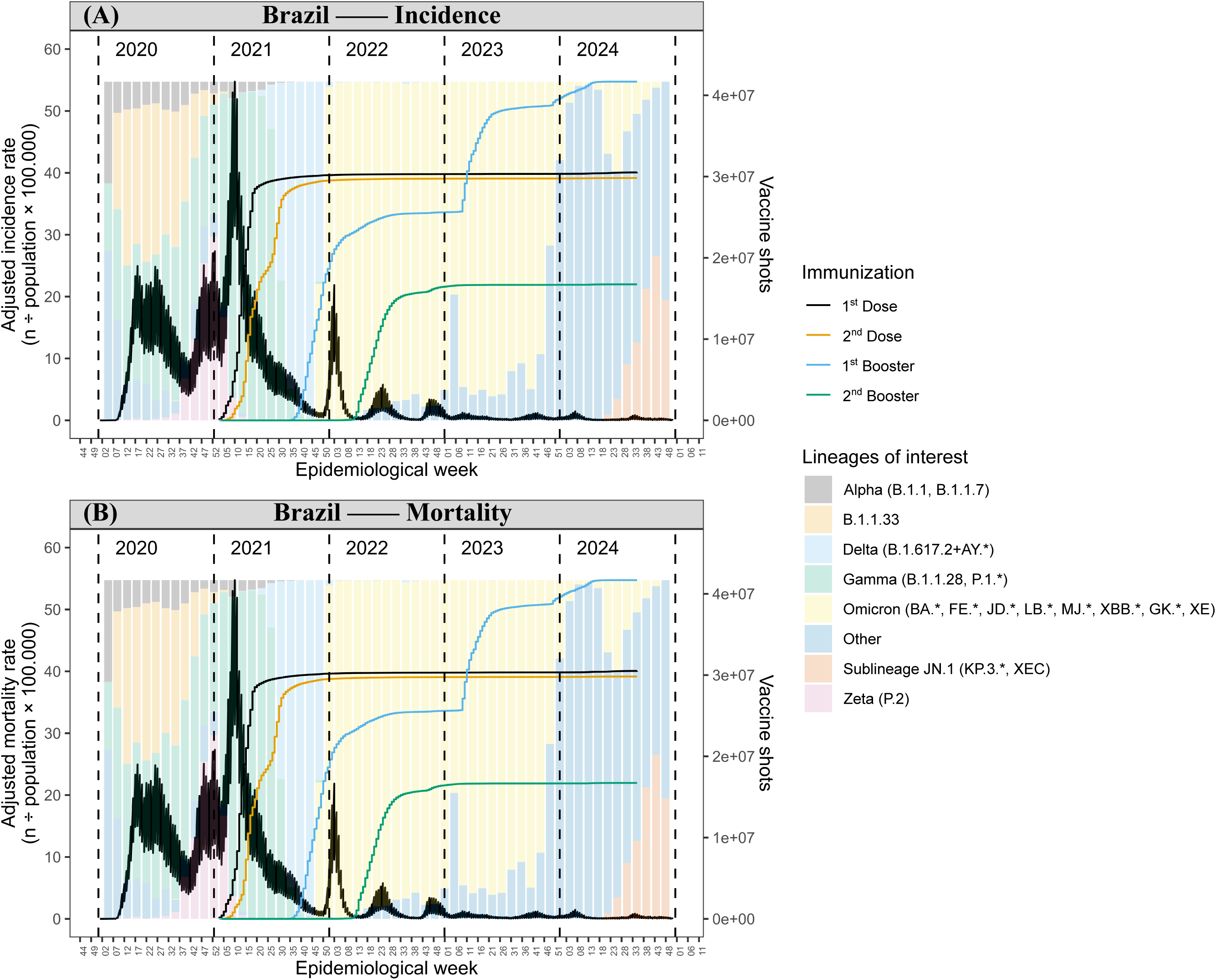
Temporal trends of adjusted incidence (A) and mortality (B) rates (grey line, left y-axis) by epidemiological week (x-axis) in Brazil from January 1, 2020, to December 31, 2024 (5 years), levels of immunization by vaccination stage (lines with right y-axis), and relative frequency of circulating variants in the territory (background bars proportional to maximum rates ranging from 0 to 100% relative frequency).

**Figure 2.**
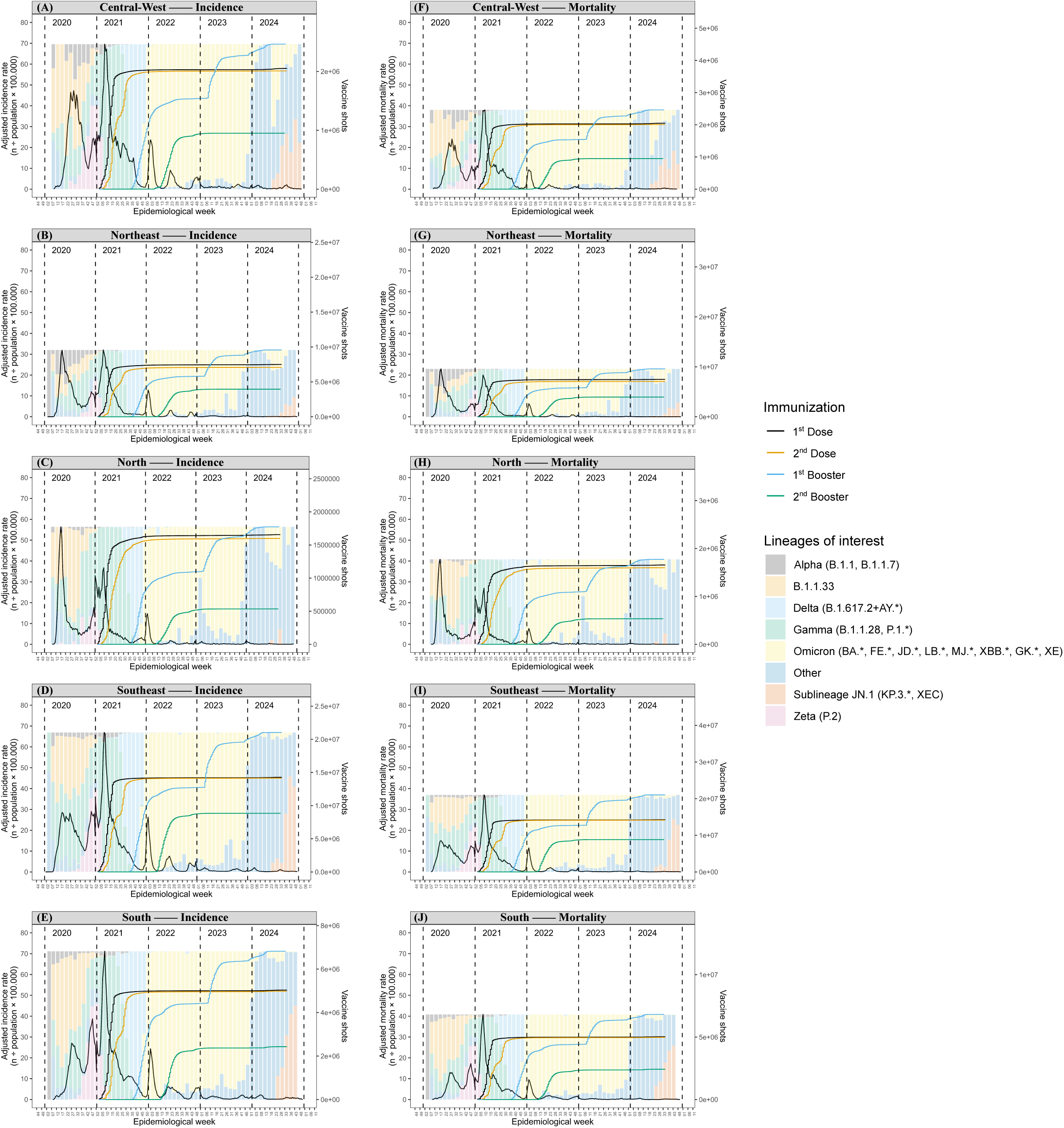
Adjusted incidence and mortality rates of SARS (left y-axis) by epidemiological week (x-axis), relative frequency of circulating variants (background bars proportional to maximum rates ranging from 0 to 100% relative frequency), and vaccination progress observed across different regions of Brazil from January 1, 2020, to December 31, 2024 (5 years). Incidence was irregular (A-E), with Omicron being the most prevalent variant. Three distinct waves were observed during the Public Health Emergency in the North and Northeast, whereas up to five or more waves were seen in the Central-West (A), Southeast (D), and especially in the South (E). The most prominent waves across all regions coincided with the circulation of the Gamma variant, highlighting it as the most impactful in terms of incidence and mortality during 2021.

The first wave was driven by the B.1.1.33 variant and the Alpha variants (B.1.1, B.1.7). The second wave was associated with the Delta (B.1.617.2+AY*) and Gamma (B.1.1.28, P.1*) variants. The third wave was dominated by the Omicron variant (BA., FE., JD., LB., MJ., XBB., GK., XE), which prevailed from 2021 through early 2024. After a subsequent decline in rates, unidentified variants emerged, and in the first quarter of 2024, the Variant of Interest (VOI) JN.1 (KP.3, XEC) arose, triggering new fluctuations in incidence and mortality rates (Figures 1-2).

During the study period, 580,818 PCR-confirmed cases of COVID-19 among individuals aged 60 to 112 years progressed to SARS, of which 304,341 resulted in recovery and 276,477 in death, corresponding to a case fatality rate (CFR) of 47.6%, with significant variation across all sociodemographic, socioeconomic, and health-related variables (χ², p<0.05) (Table 1). These variables - a total of 73 - were also fitted in univariate Generalized Linear Models (GLMs) (Table S1), and only those with p-value < 0.05 and odds ratio (OR) > 1.0 were retained for further analysis (Figure 3).

**Table 1.**
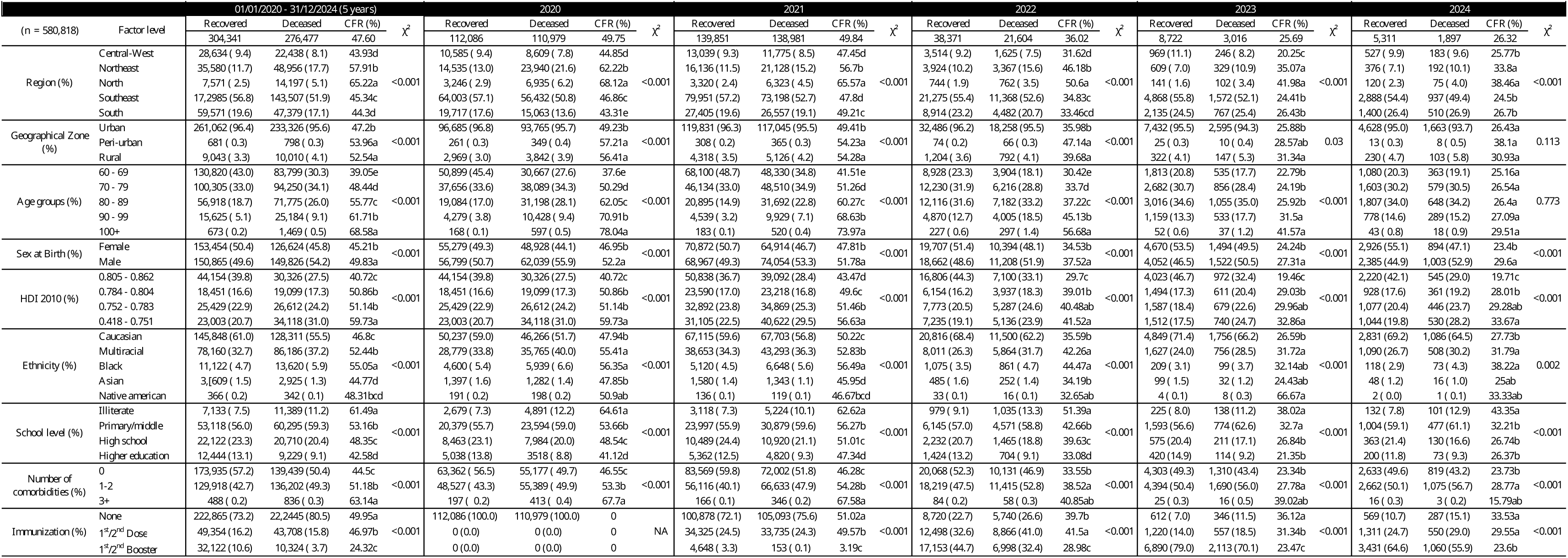
Demographic characteristics of the elderly population (60-112 years) with PCR-confirmed cases of severe acute respiratory syndrome (SARS) due to COVID-19 in Brazil, by year, from 01/01/2020 to 12/31/2024 (5 years). The χ² test indicated significant differences between the proportions of recovery and death across all strata (χ², p < 0.001). Case fatality rates (CFR%) were compared using paired Z-tests (p < 0.05). Significance letters indicate statistically significant differences between groups, with similar letters representing grouped categories, and proportions presented in descending order (from highest to lowest).

**Figure 3.**
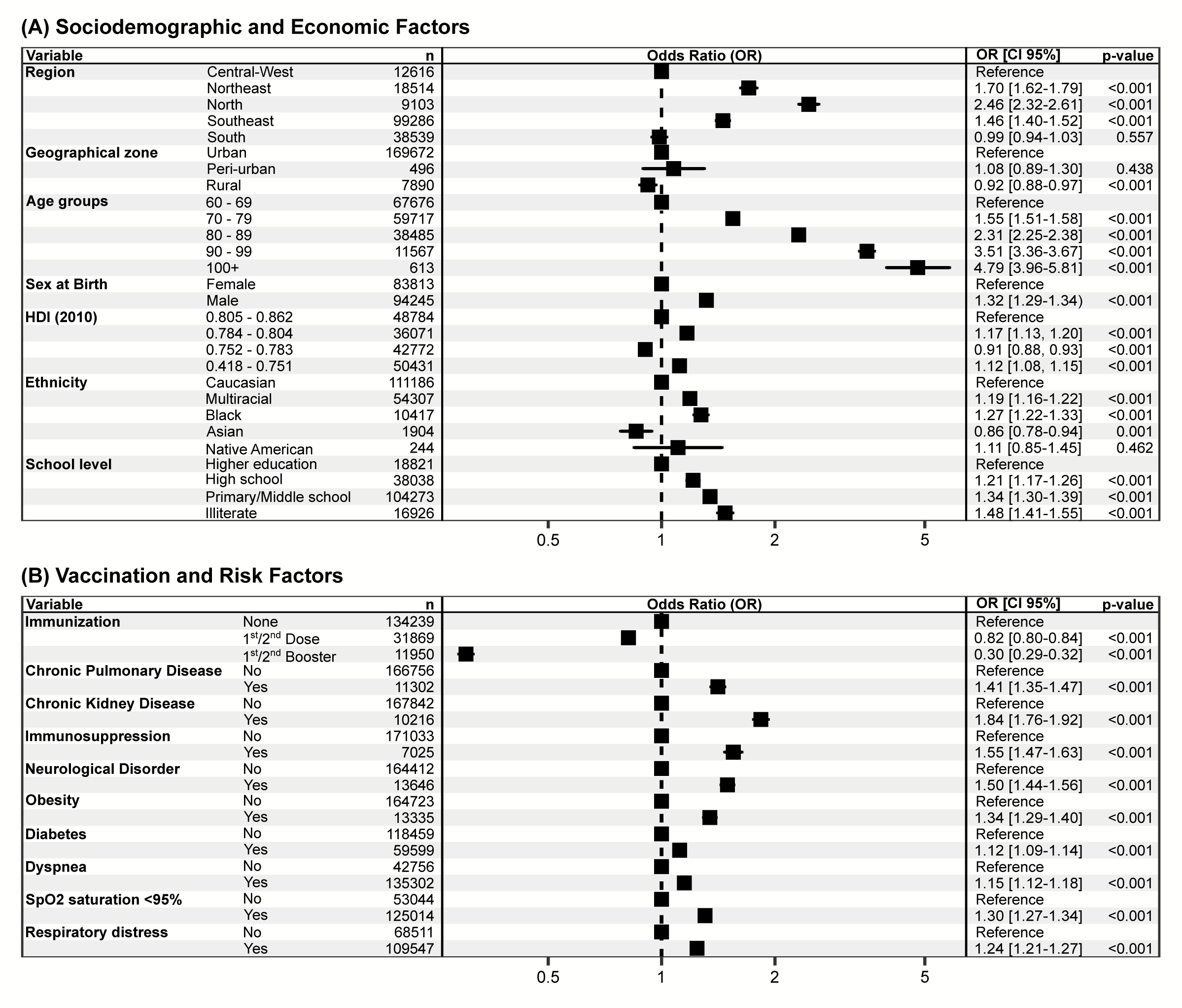
Forest plots presenting the Odds Ratios (OR) estimated from the binomial Generalized Linear Model, indicating the proportional association between independent variables and the occurrence of death. Each dot represents the OR with 95% confidence intervals, and p-values < 0.05 indicate statistical significance. The X-axis displays the OR, where ‘1’ represents the reference (no proportional effect relative to the baseline level). Panel (A) analyses sociodemographic and economic variables (region, HDI, age group, sex, ethnicity, education, and geographic zone), while panel (B) explores variables related to vaccination, as well as the presence of comorbidities, signs, and symptoms retained in the multivariate model. Both panels are part of the same adjusted model. OR values > 1 indicate higher odds of death or aggravating factors, while OR values < 1 indicate lower odds or protective factors.

### Sociodemographic and Economic Factors

Regarding case fatality rates across Brazil’s major regions, the North (CFR: 65.22%; OR: 2.46 [2.32-2.61], p<0.05) and Northeast (CFR: 57.91%; OR: 1.70 [1.62-1.79], p<0.05) regions exhibited the highest lethality, followed by the Southeast (CFR: 45.34%; OR: 1.46 [1.40-1.52], p<0.05). In contrast, the South and Central-West regions showed similar lethality rates (Z-test, p>0.05), with the South presenting a CFR of 44.30% (OR: 0.99 [0.94-1.03], p>0.05) and the Central-West 43.93% (OR: *Reference*) (Table 1, Figure 3A).

The vast majority of cases were recorded among individuals residing in urban areas (96.4% of recoveries and 95.6% of deaths). Although cases from peri-urban and rural areas represented less than 5.0% of all reported recoveries and deaths, a higher case fatality rate was observed with increasing distance from urban centre (Table 1). Individuals from peri-urban areas had the highest CFR (53.96%, OR: 1.08 [0.89-1.30], p>0.05), followed by those from rural areas (52.54%, OR: 0.92 [0.88-0.97], p<0.05). Finally, residents of urban areas, despite accounting for over 95% of all cases, had the lowest CFR (47.2%, reference group) compared to those from other zones (Table 1, Figure 3A). Nevertheless, the sample imbalance between urban and peri-urban/rural residents limited the GLM’s ability to detect statistically significant differences between these groups.

Individuals were classified into age groups as follows: 60-69 years, 70-79 years, 80-89 years, 90-99 years, and 100 years or older. Regarding age groups, there was a marked increase in case fatality rate (CFR) with advancing age (Table 1). The lowest CFR was observed in the 60-69 years group (CFR: 39.05%; OR: *Reference*), whereas the 70-79 years group showed an almost 10-point increase, with a CFR of 48.44% (OR: 1.55 [1.51-1.58], p<0.05), followed by the 80-89 years group (CFR: 55.77%, OR: 2.31 [2.25-2.38], p<0.05), the 90-99 years group (CFR: 61.71%, OR: 3.51 [3.36-3.67], p<0.05), and the group aged 100 years or older (CFR: 68.58%, OR: 4.79 [3.96-5.81], p<0.05) (Table 1, Figure 3A).

Male individuals had a higher CFR (49.83%) (OR: 1.32 [1.29-1.34], p<0.05) compared to female individuals (45.21%; OR: *Reference*) (Table 1, Figure 3A).

The 2010 Human Development Index (HDI) was used as a socioeconomic indicator, measured at the municipality level. Individuals were grouped according to HDI stratification into minimum, quartiles, and maximum ranges. A clear trend was observed: as HDI decreased, CFR increased. Individuals residing in municipalities with an HDI between 0.418 and 0.751 had a CFR of 59.73% (OR: 1.12 [1.08-1.15], p<0.05). Those residing in municipalities with an HDI between 0.752 and 0.783 had a CFR of 51.14%, showing a protective effect (OR: 0.91 [0.88-0.93], p<0.05). Subsequently, those living in areas with an HDI between 0.784 and 0.804 had a CFR of 50.86% (OR: 1.17 [1.13-1.20], p<0.05), while individuals in municipalities with an HDI between 0.805 and 0.862 had the lowest CFR (40.72%, OR: *Reference*) (Table 1, Figure 3A).

Regarding ethnicity, higher CFRs were observed among Black individuals (55.05%; OR: 1.27 [1.22-1.33], p<0.05) and Multiracial individuals (52.44%; OR: 1.19 [1.16-1.22], p<0.05), followed by Caucasian individuals (46.8%; OR: Reference) and Asian individuals (44.77%; OR: 0.86 [0.78-0.94], p<0.05). Finally, a CFR of 48.31% was observed among Native American People of Brazil. However, the Z-test and χ²-test could not detect statistical significance, likely due to sample size limitations, and the GLM model also did not identify a statistically significant association (OR: 1.11 [0.85-1.45], p>0.05), as the confidence interval ranged between OR=1 (Table 1, Figure 3A).

Educational attainment showed a gradient relationship with CFR. Individuals with higher education had the lowest CFR (42.58%; OR: *Reference*). Those with a secondary (high school) education had a CFR of 48.35% (OR: 1.21 [1.17-1.26], p<0.05), and those with primary education had a CFR of 53.16% (OR: 1.34 [1.30-1.39], p<0.05). Illiterate individuals exhibited the highest CFR and OR (61.49%; OR: 1.48 [1.41-1.55], p<0.05) (Table 1, Figure 3A).

The absence of comorbidities was associated with the lowest CFR (44.5%). The presence of one or two comorbidities increased the CFR to 51.18%, and the highest CFR was observed among individuals with three or more comorbidities (63.14%).

Regarding immunization status, the highest CFR was observed among individuals who had not received any vaccination (49.95%; OR: *Reference*), followed by those who received only the 1^st^/2^nd^ Dose (46.97%; OR: 0.82 [0.80-0.84], p<0.05). The lowest CFR was found among individuals who received the 1^st^/2^nd^ Booster, with a CFR of 24.32% (OR: 0.30 [0.29-0.32], p<0.05) (Table 1, Figure 3A).

### Sociodemographic, Economic, Risk, and Vaccination Factors by Year

When analysing CFRs by region separately for each year (2020 to 2024), it was observed that the Central-West region consistently exhibited the lowest CFRs in 2020, 2021, 2022, and 2023 (44.85%, 47.45%, 31.62%, and 25.77%, respectively). The highest CFRs in 2020, 2021, and 2022 were observed in the North region (68.12%, 65.57%, and 50.6%, respectively), whereas in 2023 and 2024, the North region (41.98% and 38.46%) presented CFRs similar to those of the Northeast region (35.07% and 33.80%) (Table 1).

Regarding CFR by age group among the elderly each year, the 60-69 age group consistently had the lowest CFRs in 2020, 2021, and 2022 (37.6%, 41.51%, and 30.42%, respectively), while individuals aged 100 years or older exhibited the highest CFRs (78.04%, 73.97%, and 56.68%, respectively). In 2023, no significant differences were observed among the 60-69 (22.79%), 70-79 (24.19%), and 80-89 (25.92%) age groups, all of which recorded the lowest CFRs that year. Meanwhile, the 90-99 (31.5%) and 100+ (41.57%) age groups had the highest CFRs, without significant differences between them. By 2024, there were no statistically significant differences between any elderly age groups (Table 1).

Regarding the influence of HDI on CFR, across all years analysed (2020 to 2024), the highest CFRs were consistently found among individuals living in municipalities with the lowest HDI strata (59.73%, 56.63%, 41.52%, 32.86%, and 33.67%, respectively), while the lowest CFRs were consistently observed among those living in municipalities with the highest HDI strata (40.72%, 43.47%, 29.7%, 19.46%, and 19.71%, respectively) (Table 1).

For sex at birth, in all years analysed (2020 to 2024), males consistently showed higher CFRs (52.2%, 51.78%, 37.52%, 27.31%, and 29.60%, respectively) compared to females (46.95%, 47.81%, 34.53%, 24.24%, and 23.4%, respectively) (Table 1).

When CFR was evaluated by ethnicity yearly (2020 to 2024), a different pattern was observed annually. In 2020, the Black (56.35%) and Multiracial (55.41%) populations exhibited the highest CFRs, with no significant difference between them. In 2021, the Black population had the highest CFR (56.49%) compared to all other groups. In 2022, both the Black (44.47%) and Multiracial (42.26%) populations again had the highest CFRs, without a significant difference between them, and were comparable to the Indigenous population (32.65%). In 2023 and 2024, there were no significant differences in CFRs among the Black (32.14% and 38.22%), Multiracial (31.72% and 31.7%), Asian (24.43% and 25.00%), and the Native American People of Brazil (66.67% and 31.79%) (Table 1).

Regarding the impact of education level on CFR, in 2020, 2021, 2022, and 2024, individuals classified as illiterate presented the highest CFRs (64.61%, 62.62%, 51.39%, and 43.35%, respectively). An exception occurred in 2023, when the highest CFRs were observed among the illiterate population (38.02%) and those with only elementary education (32.7%), with no significant difference between these groups (Table 1).

As for geographic zone, in 2020, 2021, and 2022, the highest CFRs were observed among residents of urban areas (49.23%, 49.41%, and 35.98%, respectively). In 2023, there was no significant difference between CFRs in urban (25.88%) and peri-urban (28.57%) zones, nor between peri-urban and rural zones (31.34%). By 2024, no significant differences were observed across geographic zones (Table 1).

Regarding the number of comorbidities and their influence on CFR, it was found that in 2020 and 2021, individuals with three or more comorbidities had the highest CFRs (67.7% and 67.58%, respectively). For the years 2022, 2023, and 2024, the highest CFRs were observed among individuals with ‘1-2 comorbidities’ (38.52%, 27.78%, and 28.77%, respectively) and ‘3+ comorbidities’ (40.85%, 39.02%, and 15.79%, respectively), with no significant differences compared to those with no comorbidities (33.55%, 23.34%, and 23.73%, respectively) (Table 1).

As for immunization status, there are no available data for 2020 since vaccination campaigns had not yet started. For 2021 to 2024, the lowest CFRs were observed among individuals who had received Booster doses (3.19%, 28.98%, 23.47%, and 23.60%, respectively) (Table 1).

### Impact of Vaccination on Reducing the Cumulative Risk of COVID-19 Mortality

Subsequently, the influence of the number of comorbidities and vaccination status on survival and cumulative risk in the study population was assessed using the Kaplan-Meier estimator (Figure 4A-B).

**Figure 4.**
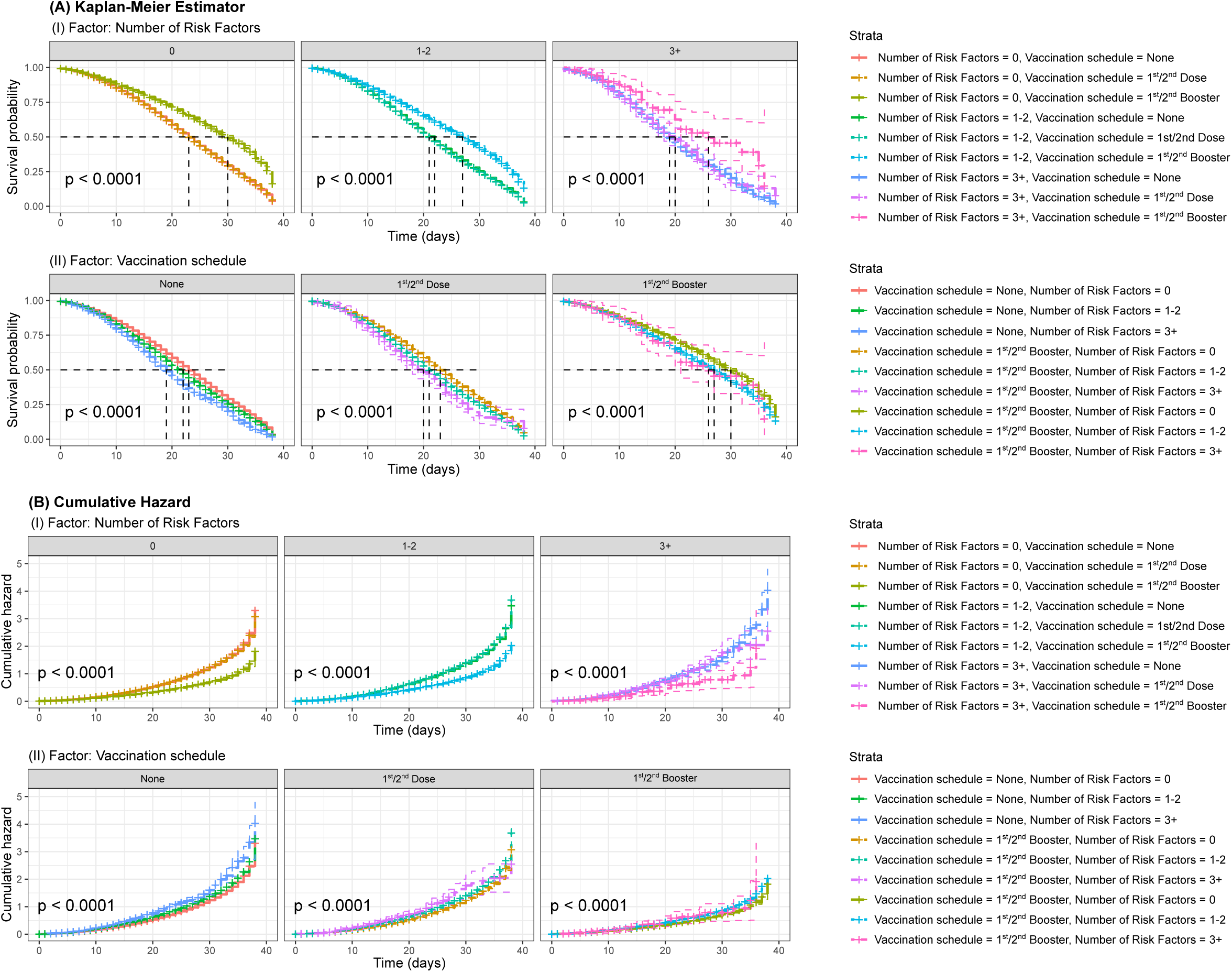
Kaplan-Meier Survival Curves (A) and Cumulative Risk of Death Curves (B) stratified by the number of comorbidities and vaccination status. In panel (A), the analysed factor is the number of comorbidities, with individuals categorized as: ‘0’ (no comorbidities), ‘1-2’ (one or two comorbidities), and ‘3+’ (three or more comorbidities). In panel (B), the assessed factor is the vaccination scheme, classified as: ‘None’ (no dose received), those immunized with one dose or the full primary series (‘1st/2nd dose’), or those immunized with the first or second booster dose (‘1st/2nd Booster’). The curves show the probability of survival over time, with p-values indicating statistically significant differences (< 0.05) between groups, reinforcing the impact of comorbidities (A) and immunization (B) on disease progression.

The vaccination schedule significantly increased the median survival time in the study population, regardless of the number of comorbidities, especially among individuals who had received the 1^st^/2^nd^ Boosters. No significant increase in survival time was observed among those who remained unvaccinated or who had only received the primary 1^st^/2^nd^ Dose schedule (Figure 4A).

When analysing the effect of immunization schedule stratified by the number of comorbidities, a noticeable reduction in survival disparities was observed, with the 1st/2nd booster group showing a ‘bulging’ of the survival curve, indicating a shift toward longer median survival (Figure 4A). In both analyses, the differences between groups were statistically significant (χ² = 2,953; dF = 8; p<0.05).

The impact of these factors on cumulative risk was also evaluated. It was observed that, regardless of the number of comorbidities, the cumulative risk of death increased over time across all groups (Figure 4B). However, unvaccinated individuals consistently exhibited a higher cumulative risk compared to those who had received, particularly, the 1^st^/2^nd^ Booster schedule (Figure 4B).

### Signs, Symptoms and Risk Factors in Cases Progressing to SARS

The risk factors retained by the GLM model (Figure 4) were chronic pulmonary disease (OR: 1.41 [1.35-1.47], p<0.05), chronic kidney disease (OR: 1.84 [1.76-1.92], p<0.05), immunosuppression (OR: 1.55 [1.47-1.63], p<0.05), neurological disorder (OR: 1.50 [1.44-1.46], p<0.05), obesity (OR: 1.34 [1.29-1.40], p<0.05), and diabetes (OR: 1.12 [1.09-1.14], p<0.05).

The signs and symptoms retained by the GLM model (Figure 4) were dyspnea (OR: 1.15 [1.12-1.18], p<0.05), oxygen saturation (SpO₂) < 95% (OR: 1.30 [1.27-1.34], p<0.05), and respiratory distress (OR: 1.24 [1.21-1.27], p<0.05).

## DISCUSSION

This study explores the dynamics of COVID-19 cases among the elderly population in Brazil between January 2020 and December 2024, spanning five years and encompassing both mild and moderate cases reported through Influenza-like Illness surveillance (ILI) and severe cases requiring hospitalization for Severe Acute Respiratory Syndrome (SARS). The analysis integrates interconnected factors such as the emergence of new variants, the vaccination campaign, the circulation of SARS-CoV-2 variants across Brazil, and the demographic and clinical characteristics of the susceptible population.

When comparing laboratory-confirmed cases of Influenza-like Illness (ILI) and Severe Acute Respiratory Syndrome (SARS) across age groups, a striking inversion of patterns emerged. Among the elderly, ILI was relatively uncommon (n=15,658), whereas SARS predominated with 580,818 cases. In this age group, ILI accounted for only 2.7% of confirmed cases, while SARS represented nearly half of all PCR-confirmed SARS cases nationwide. In contrast, the pattern was reversed in younger populations. Among children aged 0 to 19 years (unpublished data), the vast majority of confirmed cases were ILI (n=2,680,871), with only 38,975 SARS cases, meaning that ILI constituted 98.6% of laboratory-confirmed cases in this group. These contrasting trends clearly illustrate that ILI was predominantly observed in younger individuals, whereas SARS disproportionately affected older adults.

### The Role of Emerging Variants in Shaping Pandemic Waves and Disease Severity

SARS-CoV-2 variants developed apomorphies that enhanced their transmissibility and lethality. These variants accumulated genetic modifications that resulted in amino acid substitutions, primarily in the gene responsible for encoding the Spike protein, which mediates interaction with the host cell’s ACE2 receptor. Such changes affected various aspects of pathogenicity across different variants^28^.

At the onset of the pandemic in Brazil, the VOI B.1.1.33 and VOCs Gamma (B.1.1.28, P.1.*) and Omicron (BA.*, FE.*, JD.*, LB.*, MJ.*, XBB.*, GK.*, XE) were the predominant circulating variants (Figures 2-3). These variants already carried the E484K mutation, associated with reduced neutralization by antibodies generated either through previous infection or vaccination^28^. This likely contributed to sustaining the waves of cases via reinfections occurring within short time intervals^29^, particularly due to their rapid mutational capacity, which facilitated the emergence of new lineages^30^.

VOC Gamma was primarily responsible for the second wave in Brazil, which recorded the highest lethality during the pandemic period, marking a critical point that coincided with the early stages of vaccination rollout (Figures 2-3). Subsequently, VOC Delta (B.1.617.2+AY.) became predominant but had a relatively attenuated impact compared to its predecessors^31^. The end of the pandemic period was marked by the dominance of VOC Omicron, which triggered its own wave and remained the most prevalent variant from late 2021 through 2023, continuing to circulate even after the pandemic emergency had ended. Despite being less pathogenic than VOCs Gamma or Delta, Omicron exhibited robust immune and vaccine escape mechanisms combined with high transmissibility. These characteristics highlighted the necessity of completing booster vaccination schedules to reduce the risk of progression to SARS^32^.

### Sociodemographic and economic Factors Shaped COVID-19 Outcomes in Older Adults

Regional disparities became evident when analysing the evolution of incidence and mortality rates across different territories. Although the North and Northeast presented the lowest adjusted incidence and mortality rates (Figure 3), they recorded the highest CFRs when compared to other regions (Tables 1 and 2). This outcome was linked to limited access to and quality of healthcare services. The vast geographic area of the North contributes to population dispersion, hindering access to hospitals and forcing residents of remote areas to travel to capital cities for specialized medical care^33^. This issue is similar in the Northeast, where socioeconomic factors such as poverty, inequality, and high rates of informal labour further intensified the challenges^34^.

Higher fatality rates among individuals aged 60 years and older were consistent with previous studies^35–37^, reflected on the effects of comorbidities/risk factors and declining immune function in this age group^38,39^. Increased CFR among males was also observed and may be associated with sex-based differences in comorbidities^36,40,41^.

Among the social determinants of health, including social, political, environmental, economic, and cultural factors affecting healthcare access, those linked to income distribution were identified as having the greatest impact on quality of life and population health^42^.

With regard to ethnicity, Black and Multiracial individuals experienced higher CFRs compared to the Caucasian population, consistent with other studies highlighting increased COVID-19 lethality in these ethnic groups^43,44^. This discrepancy is a globally documented phenomenon. For example, in the United States, Hispanic populations were most affected, followed by Indigenous peoples, Asians, Black and Alaska Natives People, while Caucasian had the lowest mortality rates^43^. Such differences are thought to reflect structural socioeconomic inequalities and historical disparities that limit social mobility among racial groups. In Brazil, these inequities manifest in restricted healthcare access and broader social and economic disadvantages, disproportionately affecting Black and Multiracial populations^45,46^.

Moreover, higher education levels were associated with lower mortality rates, a finding consistent with studies identifying education, alongside socioeconomic factors, as a key determinant of access to healthcare^44,47–49^. Low educational attainment is directly linked to higher rates of informal employment^50^, a condition that increases social vulnerability. The absence of labour protections for informal workers prevented many from adhering to social distancing measures during the pandemic, which in turn contributed to prolonged exposure to SARS-CoV-2 and increased incidence and mortality in these groups.

### Ageing and multimorbidity

Since the beginning of the pandemic, studies have indicated that individuals with chronic diseases or comorbidities/risk factors experienced more severe outcomes, which helped identify and prioritize risk groups for medical interventions and vaccination campaigns.

Multimorbidity is directly associated with aging, significantly altering health profiles from the age of 60 onwards^51^. Although establishing direct causal links between comorbidities and COVID-19 severity remains challenging^52^, higher CFRs, shorter median survival times and increased cumulative risk were observed in individuals with a greater number of comorbidities (Table 1, Figure 4). The worsening of COVID-19 cases has been linked to a hyperinflammatory syndrome driven by dysregulation of the renin-angiotensin system, which leads to endothelial, vascular, and immune dysfunction^53^, compromising other systems, particularly the cardiovascular and renal systems^54^.

Within the studied population, the comorbidities most strongly associated with unfavourable outcomes included chronic lung disease, kidney disease, immunosuppression, neurological disorders, obesity, and diabetes. Clinical signs such as dyspnea, reduced oxygen saturation, and respiratory distress were also important predictors of severity (Figure 3).

Alongside the increasing number of comorbidities, aging also brings biological changes to the immune system that impair disease response and increase susceptibility to infections, while also influencing clinical progression and long-term sequelae after recovery^6^.

Survival curves stratified by number of comorbidities (Figure 4) demonstrate a negative association between comorbidities/risk factors and median survival time. Individuals without comorbidities had higher probabilities of survival over time, whereas those with one or more comorbidities experienced progressively reduced survival. The group with three or more comorbidities (‘3+’) showed the steepest decline in survival probability, regardless of vaccination status. The differences between groups were statistically significant (p<0.05), highlighting the detrimental impact of multimorbidity on survival.

Cumulative risk of death curves (Figure 4) further reinforces the negative influence of multimorbidity on survival. Individuals with a higher burden of comorbidities accumulated risk of death more rapidly over time. The group with three or more comorbidities displayed a steeper increase in cumulative risk, indicating a rising mortality rate as time progressed. In contrast, individuals without comorbidities exhibited significantly lower cumulative risk. These findings underscore multimorbidity as a key determinant of poorer prognosis, regardless of vaccination status (p<0.05).

### The Protective Role of Vaccination and Boosters in Reducing COVID-19 Lethality

Studies have shown that vaccines demonstrated a significant increase in efficacy after the completion of the vaccination schedule, with the protective effect becoming evident 14 days after administration, regardless of the technology or platform used for their development^55–58^.

Vaccination had a positive impact on reducing lethality rates, particularly following the administration of booster doses (Figures 3-4). Unvaccinated individuals accounted for a large portion of those at risk and who ultimately died, showing significantly higher CFRs (Table 1). Despite progress in the immunization campaign, vaccination coverage remained uneven, especially when considering the number of doses administered up to the 2nd booster dose. Moreover, the effectiveness of vaccines was somewhat limited in the context of the rapid emergence and spread of SARS-CoV-2 variants.

## CONCLUSIONS

SARS cases in the elderly accounted for half of all SARS cases confirmed by PCR in Brazil. The management of COVID-19 in the country highlighted deep-rooted structural and social issues. In addition to the likely predisposition of the elderly population to progress to SARS, driven by factors such as immunosenescence and the aggravating interaction between multimorbidity and pathophysiological mechanisms, there was also a clear socioeconomic profile among the most affected. These predominantly included Black and Multiracial individuals living in the North and Northeast regions, as well as rural and peri-urban areas, where there are socioeconomic inequalities and access to healthcare services.

Alongside the factors inherent to the study population, the rapid emergence and spread of new variants with immune escape capabilities may have been a critical driver for the persistence of the pandemic scenario.

Both multimorbidity and lack of vaccination were associated with poorer prognoses, reflected in reduced survival and higher cumulative mortality risks. The interplay between these factors underscores the urgent need for prevention strategies, including expanding vaccination coverage, particularly among populations with higher numbers of comorbidities.

The findings reinforce the necessity of strengthening and decentralizing health systems away from metropolitan areas, broadening access to immunization, and implementing continuous monitoring strategies to improve preparedness for future public health emergencies.

## Data Availability

All bash and R script/codes will be released on GitHub's Corresponding Author upon publishing of the material.

## ACKNOWLEDGEMENTS

The authors thank the Brazilian Government and the Brazilian Ministry of Health for making public secondary data available and acknowledge all individuals directly and indirectly involved in the efforts to manage the COVID-19 crisis in the country.

Silvio Alencar Cândido-Sobrinho gratefully acknowledges the Coordination for the Improvement of Higher Education Personnel (CAPES/MEC/BRAZIL) for the study grant (grant number: 88887.479534/2020-00) - Finance Code 001.

## CONFLICT OF INTEREST

The authors declare no competing interest.

## DATA SHARING

Bash and R scripts used in this research will be made available at the Corresponding Author’s GitHub upon publishing in a journal.

---

**Figure S1.**
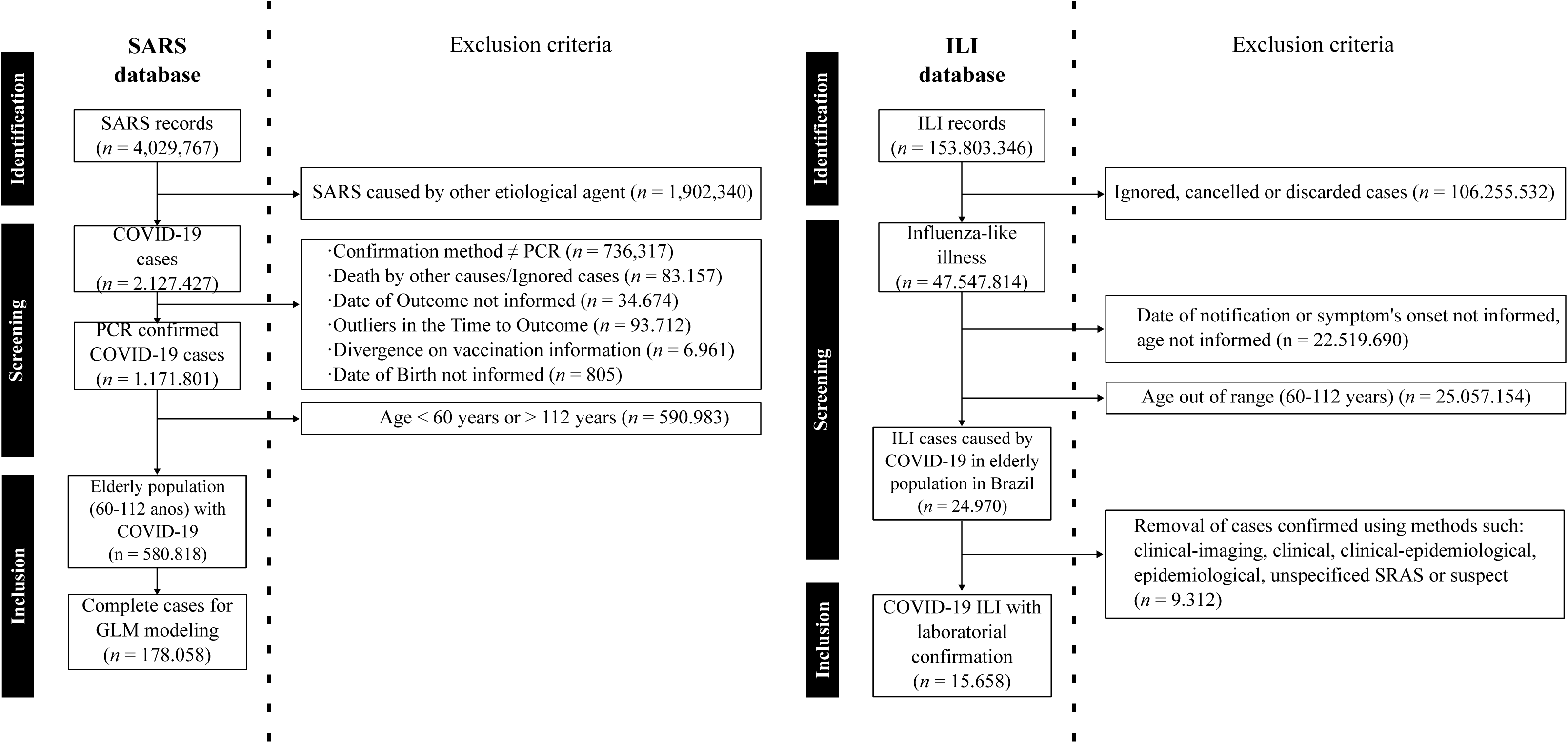
Flowchart of the inclusion and exclusion criteria for elderly patients (≥60 and ≤112 years) with mild and moderate cases (ILI) and severe cases (SARS) of COVID-19 in Brazil, from 01/01/2020 to 12/31/2024 (5 years). Cases were excluded based on the following criteria: infections by etiological agents other than SARS-CoV-2, confirmation methods other than PCR, death due to causes unrelated to COVID-19, missing outcome data (ignored, cancelled, or discarded), and inconsistencies in the records (such as unidentified age, discrepant discharge dates, or missing data).

## Notes

### Competing Interest Statement

The authors have declared no competing interest.

### Funding Statement

This study was funded by Coordination for the Improvement of Higher Education Personnel (CAPES/Ministry of Education/Brazil) by the grant number: 88887.479534/2020-00, Finance Code 001.

### Author Declarations

Dear referee, This study use secondary data released by the Brazilian Ministry of Health and affiliated institutes. The data available is anonymized, compliant with the Brazilian framework for personal data protection ('LGPD') released by OpenDataSus, a branch from the Ministry of Health. The data is released under license CC-BY. Data from all ILI and SARS cases registered in Brazil may be retrieved from here (link:https://opendatasus.saude.gov.br/en/dataset/?q=síndrome) Data for the Brazilian National Vaccination Schedule/Campaign may be retrieved from here (link:https://opendatasus.saude.gov.br/en/dataset/covid-19-vacinacao) Data from Genomic Surveillance reported by territories, not possible to identify an individual may be retrieved from here (link:https://opendatasus.saude.gov.br/en/dataset/covid-19-vacinacao)

### Summary of Updates

Figures 1 and 2 revised; text was revised to match up with figures information.

